# Sex-specific DNA methylation changes in Alzheimer’s disease pathology

**DOI:** 10.1101/2021.03.01.21252029

**Authors:** Lanyu Zhang, Juan I. Young, Lissette Gomez, Tiago C. Silva, Michael A. Schmidt, Jesse Cai, Xi Chen, Eden R. Martin, Lily Wang

## Abstract

Sex is an important factor that contributes to the clinical and biological heterogeneities in Alzheimer’s disease (AD), but the regulatory mechanisms underlying sex disparity in AD are still not well understood. DNA methylation is an important epigenetic modification that regulates gene transcription and is known to be involved in AD. We performed the first large-scale sex-specific meta-analysis of DNA methylation changes in AD, by re-analyzing four recent epigenome-wide association studies totaling more than 1000 postmortem prefrontal cortex brain samples using a uniform analytical pipeline. For each cohort we employed two complementary analytical strategies, a sex-stratified analysis that examined methylation-Braak stage associations in male and female samples separately, and a sex-by-Braak stage interaction analysis that compared the magnitude of these associations between different sexes. Our analysis uncovered 14 novel CpGs, mapped to genes such as *TMEM39A* and *TNXB* that are associated with AD in a sex-specific manner. *TMEM39A* is known to be involved in inflammation, dysregulated type I interferon responses, and other immune processes. *TNXB* encodes tenascin proteins, which are extracellular matrix glycoproteins demonstrated to modulate synaptic plasticity in the brain. Moreover, for many previously implicated AD genes, such as *MBP* and *AZU1*, our analysis provided the new insights that they were predominately driven by effects in only one sex. These sex-specific DNA methylation changes were enriched in divergent biological processes such as integrin activation in females and complement activation in males. Importantly, a number of drugs commonly prescribed for AD patients also targeted these genes with sex-specific DNA methylation changes. Our study implicated multiple new loci and biological processes that affected AD in a sex-specific manner and highlighted the importance of sex-specific treatment regimens for AD patients.

## Introduction

Alzheimer’s disease (AD) is the most common cause of dementia. With the aging population in the U.S., AD has become a major public health concern and one of the most financially costly diseases [1]. Almost two-thirds of AD patients in the U.S. are women [2]. After diagnosis, women also progress faster with more rapid cognitive and functional declines [3–8]. On the other hand, it has also been reported men with AD have increased risk for death [9–11]. However, the molecular mechanisms underlying these observed disparities in AD are still not well understood. Previous studies have shown that epigenetics is an important contributor to the sex differences in brain functions and vulnerability to diseases [12–16]. Among epigenetic modifications, DNA methylation profiles differ significantly between males and females at a number of loci in adult brains [17]. Importantly, alterations of DNA methylation levels have also been implicated in multiple neurological disorders including AD [18–22].

However, thus far, a comprehensive characterization for the contribution of sex to DNA methylation changes in AD has not been performed. In the identification of sex-specific effects, statistical power is a major challenge [23]. Stratifying by sex reduces the sample size of both groups. Also, comparing methylation to disease associations between the sexes by testing interaction effect would require a much larger sample size than detecting a main effect with the same magnitude [24]. To address these challenges, we performed a comprehensive meta-analysis of more than 1000 post-mortem brain prefrontal cortex samples, collected from four recent AD epigenome-wide association studies [18–21], to identify the most consistent DNA methylation changes affected by AD in a sex-specific manner. As sex is a strong factor in driving inter-personal variabilities in AD, the results of this study are particularly relevant for precision medicine.

## Methods

### Study cohorts

Our meta-analysis included 1,030 prefrontal cortex brain samples (642 female samples and 388 male samples) from four independent cohorts (Supplementary Table 1), previously described in the ROSMAP [18], Mt. Sinai [20], London [19], and Gasparoni [21] DNA methylation studies.

### Pre-processing of DNA methylation data

As described elsewhere [22], for each cohort quality control included removing probes with detection P-value < 0.01 in all samples and those associated with cigarette smoking [25] or SNPs, and removing samples with low bisulfite conversion efficiency or detected as outliers in principal component analysis (see details in Supplementary Note 1). Next, the QN.BMIQ normalization procedure [26] was performed on the quality-controlled methylation data, followed by fitting linear model `methylation M value ∼ methylation slide` to remove batch effects. The `methylation residuals` from these linear models were then used for subsequent analysis.

### Single cohort and meta-analysis

In sex-stratified analysis, for each CpG, we applied the model methylation residuals ∼ age at death + Braak stage + CETS estimated neuron proportions [27] to female samples and male samples separately. For the analysis of differentially methylated regions (DMRs), we used the coMethDMR R package [28] to identify co-methylated DMRs associated with Braak stage (Supplementary Note 2), by implementing the same linear model described above. We considered CpGs (or DMRs) with false discovery rate (FDR) less than 0.05 in female samples or male samples to be significant.

To assess inflation of the test statistics, we used quantile-quantile (QQ) plots and estimated genomic inflation factors using both the conventional approach and the *bacon* method [29] (Supplementary Note 3). The *bacon* method was also used to obtain inflation-corrected effect sizes, standard errors, and p-values for each cohort, which were then combined by inverse-variance weighted meta-analysis models using R package meta (Supplementary Note 4).

In sex-by-Braak stage interaction analysis, for each CpG, we applied the model methylation residuals ∼ age at death + sex + Braak stage + sex*Braak stage + sex*age at death + CETS estimated neuron proportions to samples including both sexes. To select significant CpGs and DMRs, we applied a stagewise analysis approach, previously proposed by van de Berge et al. (2017) [30] (Supplementary Note 5), which was shown to have improved power in high-throughput experiments where multiple hypotheses are tested for each gene.

### Enrichment and pathway analysis

We tested over- and under-representation of significant CpGs and DMRs in different types of genomic regions and chromatin states using Fisher’s exact test. Pathway analysis was performed by comparing the genes with significant sex-specific DNA methylation changes in AD with the canonical pathways and biological process GO terms in MSigDB using GSEA analysis [31] (Supplementary Note 6).

### Integrative methylation – gene expression analysis

Integrative methylation – gene expression analysis was performed using the ROSMAP study samples with matched DNA methylation and gene expression data. First, we linked significant CpGs (or DMRs) to nearby genes using GREAT [32]. Next, we removed confounding effects due to batch, age at death, and cell types in methylation data and gene expression data separately by fitting linear models and extracting residuals. Finally, for each gene expression – CpG (or DMR) pair, we tested the association between gene expression residuals and methylation residuals, adjusting for Braak stage (Supplementary Note 7).

### Sex-specific mQTL analysis

The ROSMAP dataset, imputed to HRC r1.1 reference panel [33], with matched genotype data and DNA methylation data for 688 samples (434 females, 254 males) was used for this analysis. We considered SNPs that are within 500kb from FDR significant CpGs (or DMRs), with MAF > 1%, info score ≥ 0.4, and are significantly associated with AD status (P < 0.05) (Supplementary Note 8). Association between methylation residuals (after removing batch, age at death, and cell type effects) and SNPs were then tested using linear models, adjusting for batch in genotype data and first three PCs estimated from genotype data.

### Drug target analysis

We compared our list of sex-specific DNA methylation changes with targets of drugs prescribed to AD patients or in the development in the ChEMBL database [34] (https://www.ebi.ac.uk/chembl/). To this end, we overlapped genes mapped to significant CpGs or DMRs with the genes targeted by compounds annotated to “Alzheimer Disease” in ChEMBL.

## Results

### Description of EWAS cohorts and data

Among the four cohorts (Supplementary Table 1), the mean age at death ranged from 79.3 to 87.2 years in females and from 67.5 to 85.0 years in males. The number of CpGs and samples removed at each quality control step are presented in Supplementary Table 2. For females, inflation factor lambdas (*λ*) by the conventional approach ranged from 1.060 to 1.154, and lambdas based on the *bacon* approach [29] (*λ_bacon_*) ranged from 1.021 to 1.059 (Supplementary Figure 1). Similarly, for males, *λ* ranged from 0.906 to 1.265, and *λ_bacon_* ranged from 0.957 to 1.114. These values are comparable to those obtained in other recent large scale EWAS [35].

### Sex-specific DNA methylation changes in AD

In sex-stratified analysis, our meta-analysis identified 381 and 76 CpGs, mapped to 245 and 51 genes at 5% FDR in female and male samples, respectively (Figure 1, Table 1, Supplementary Table 3-4). Similarly, we identified 72 and 27 DMRs, mapped to 66 and 22 genes, at 5% FDR in female and male samples, respectively (Table 2, Supplementary Tables 5-6). Among them, 3.6% (16 out of 441 unique FDR significant CpGs) and 12.5% (11 out of 88 unique FDR significant DMRs) were significant in both females and males with the same direction of change. The average number of CpGs per DMR was 6.5 ± 8.9. The FDR significant methylation changes at CpGs and DMRs did not completely overlap. Only 89 out of the 381 (23.4%) significant CpGs in females, and 13 out of the 76 (17.1%) significant CpGs in males overlapped with the significant DMRs. Among all CpGs and all DMRs, the effect estimates in males and females correlated only modestly (*r*_CpG_ = 0.124, *r*_DMR_ = 0.170) and about half (53% of CpGs, 54% of DMRs) were in the same direction of change in males and females, similar to what would be expected by chance.

**Figure 1.**
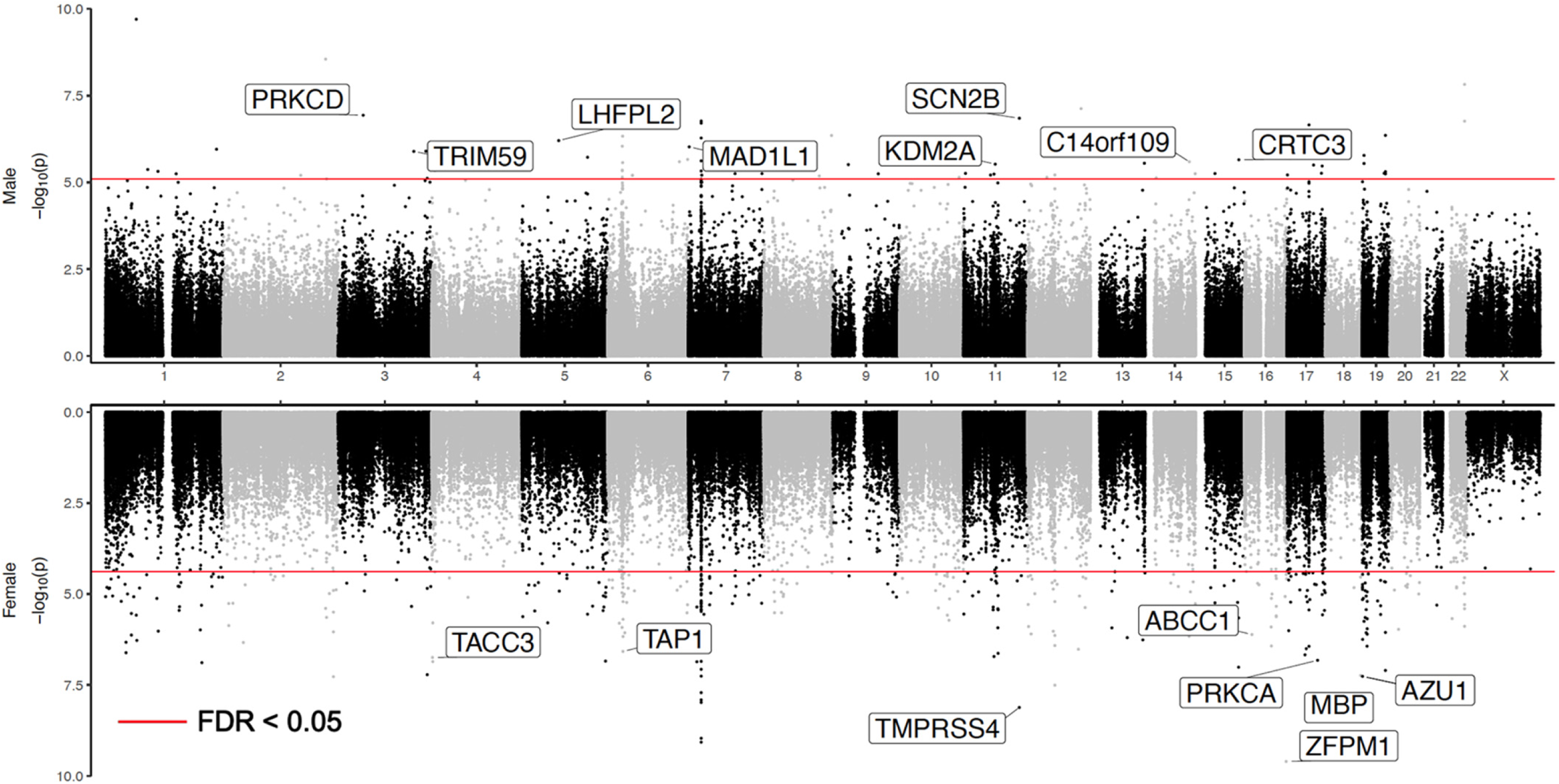
Miami plot of sex-stratified analysis results. The X-axis are chromosome numbers. The Y-axis show –log_10_(P-value) of CpG – Braak stage associations in males (above X-axis) or in females (below X-axis). The genes corresponding to top 10 CpGs that are the most significant in one sex (FDR < 0.05), but not significant in another sex (P-value > 0.05) are highlighted.

**Table 1.** Top 10 CpGs in sex-stratified analysis. Shown are CpGs that are highly significant in one sex (FDR < 0.05), but not significant (P-value > 0.05) in the other sex. For each CpG, annotations include the location of the CpG based on hg19/GRCh37 genomic annotation (chr, position), Illumina gene annotations, nearby genes based on GREAT, and chromatin state. The inverse-variance weighted meta-analysis regression models results include estimated effect size (estimate) where CpGs that are hyper-methylated in AD have positive values, and its associated standard error (se), P-value, and false discovery rate (FDR) for multiple comparison corrections. Bold indicates significant association at 5% FDR.

| cpg | Annotations |  |  |  |  | Female samples |  |  |  | Males samples |  |  |  |
| --- | --- | --- | --- | --- | --- | --- | --- | --- | --- | --- | --- | --- | --- |
|  | chr | position | Illumina | GREAT (distance to TSS) | chromatin state | estimate | se | P-value | FDR | estimate | se | P-value | FDR |
| *significant only in female samples* |  |  |  |  |  |  |  |  |  |  |  |  |  |
| cg09502865 | chr16 | 88600155 | ZFPM1 | ZC3H18(-36633); ZFPM1(+80431) | Active TSS | 0.179 | 0.028 | **2.50E-10** | **1.15E-04** | 0.094 | 0.034 | 4.46E-01 | 9.64E-01 |
| cg05235171 | chr11 | 1.18E+08 | TMPRSS4 | TMPRSS4(+10301); SCN4B(+6545) | Quiescent/Low | -0.168 | 0.029 | **7.61E-09** | **6.99E-04** | -0.025 | 0.033 | 4.47E-01 | 9.64E-01 |
| cg15610437 | chr19 | 827821 | AZU1 | AZU1(-4) | Weak Repressed PolyCo | -0.155 | 0.029 | **5.45E-08** | **1.99E-03** | -0.090 | 0.034 | 9.79E-02 | 8.25E-01 |
| cg13572782 | chr18 | 74799495 | MBP | MBP(+45229); ZNF236(+263380) | Active TSS | 0.157 | 0.029 | **5.75E-08** | **1.99E-03** | 0.047 | 0.036 | 1.90E-01 | 8.92E-01 |
| cg17881200 | chr7 | 27138850 |  | HOXA1(-3258) | Repressed PolyComb | 0.152 | 0.029 | **1.41E-07** | **2.98E-03** | 0.131 | 0.034 | 6.22E-02 | 7.73E-01 |
| cg22632947 | chr17 | 64787784 | PRKCA | CACNG5(-43450); PRKCA(+48884) | Flanking Active TSS | -0.139 | 0.026 | **1.50E-07** | **3.00E-03** | -0.005 | 0.028 | 8.57E-01 | 9.95E-01 |
| cg15467503 | chr4 | 1727234 | TACC3 | TMEM129(-4177); TACC3(+3969) | Weak transcription | -0.146 | 0.028 | **1.81E-07** | **3.45E-03** | -0.083 | 0.034 | 1.33E-01 | 8.59E-01 |
| cg26033526 | chr6 | 32819858 | TAP1 | PSMB9(-2079); TAP1(+1896) | Genic enhancers | 0.148 | 0.029 | **2.64E-07** | **4.18E-03** | -0.021 | 0.034 | 5.23E-01 | 9.73E-01 |
| cg08363067 | chr16 | 16170085 | ABCC1 | ABCC1(+126652); ABCC6(+14723) | Weak transcription | -0.120 | 0.024 | **7.68E-07** | **7.32E-03** | -0.050 | 0.028 | 7.41E-02 | 7.94E-01 |
| cg12926693 | chr6 | 36665611 |  | RAB44(-17644); CDKN1A(+19125) | Quiescent/Low | -0.149 | 0.030 | **8.59E-07** | **7.32E-03** | -0.067 | 0.035 | 5.19E-02 | 7.52E-01 |
| *significant only in male samples* |  |  |  |  |  |  |  |  |  |  |  |  |  |
| cg07687398 | chr3 | 53198666 | PRKCD | PRKCD(+3531); TKT(+91371) | Weak transcription | 0.052 | 0.030 | 8.63E-02 | 8.19E-01 | 0.183 | 0.035 | **1.16E-07** | **9.88E-03** |
| cg10513118 | chr11 | 1.18E+08 | SCN2B | SCN2B(+184) | Active TSS | 0.038 | 0.030 | 2.01E-01 | 9.17E-01 | 0.172 | 0.033 | **1.42E-07** | **9.88E-03** |
| cg21253952 | chr8 | 1.44E+08 |  | ARC(+33833); BAI1(+132209) | Weak Repressed PolyCo | -0.019 | 0.030 | 5.33E-01 | 9.84E-01 | 0.169 | 0.034 | **4.44E-07** | **1.53E-02** |
| cg18281939 | chr5 | 77783895 | LHFPL2 | SCAMP1(+127557); LHFPL2(+160) | Weak transcription | -0.022 | 0.029 | 4.55E-01 | 9.76E-01 | -0.179 | 0.036 | **6.26E-07** | **1.79E-02** |
| cg11809272 | chr6 | 31409361 |  | HLA-B(-84398); MICB(-56530) | Quiescent/Low | -0.001 | 0.031 | 9.77E-01 | 1.00E+00 | 0.161 | 0.033 | **9.08E-07** | **2.17E-02** |
| cg15952933 | chr7 | 1899886 | MAD1L1 | ELFN1(+172132); MAD1L1(+3725) | Weak transcription | -0.058 | 0.033 | 7.56E-02 | 8.00E-01 | -0.156 | 0.032 | **9.48E-07** | **2.17E-02** |
| cg11614451 | chr3 | 1.6E+08 | TRIM59 | ENSG00000248710(-113); TRIM59 | Flanking Bivalent TSS/En | -0.041 | 0.029 | 1.56E-01 | 8.92E-01 | -0.150 | 0.031 | **1.28E-06** | **2.44E-02** |
| cg18942110 | chr15 | 91072797 | CRTC3 | CRTC3(-502) | Active TSS | 0.031 | 0.030 | 3.06E-01 | 9.52E-01 | -0.164 | 0.035 | **2.23E-06** | **3.19E-02** |
| cg01655008 | chr14 | 93652954 | C14orf10 | MOAP1(-1687); TMEM251(+159) | Transcr. at gene 5' and 3' | -0.042 | 0.029 | 1.51E-01 | 8.89E-01 | -0.152 | 0.032 | **2.54E-06** | **3.33E-02** |
| cg02331272 | chr11 | 66964208 | KDM2A | ADRBK1(-69672); KDM2A(+7705) | Weak transcription | 0.058 | 0.030 | 1.10E-01 | 8.52E-01 | -0.166 | 0.036 | **2.99E-06** | **3.52E-02** |

**Table 2.**
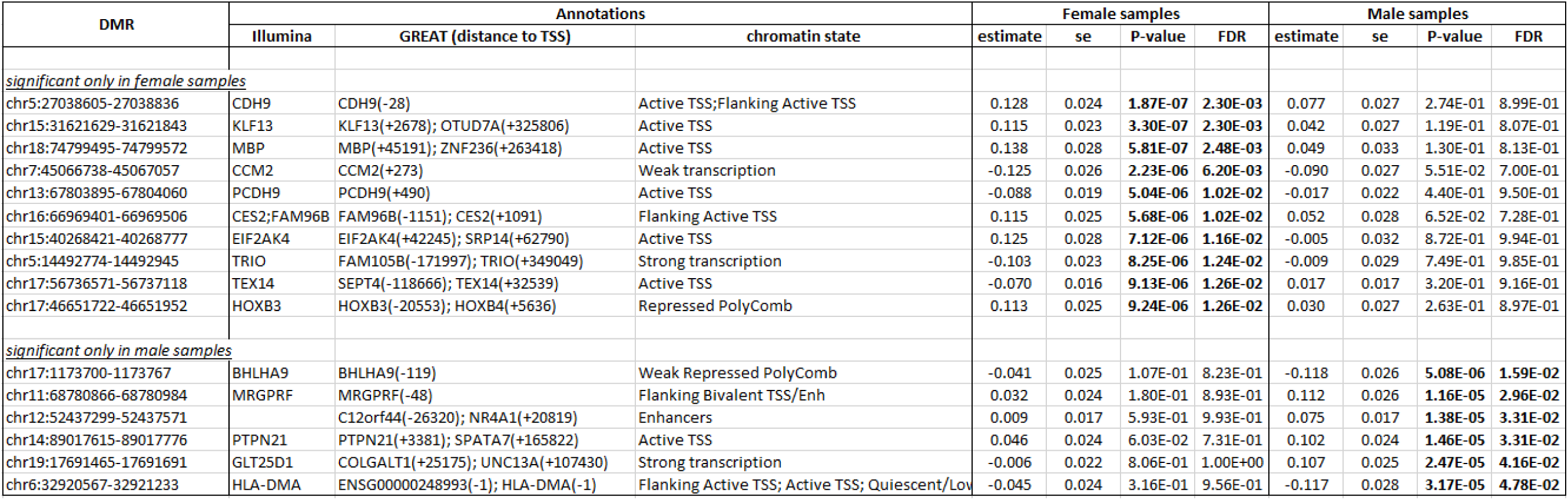
Top 10 and 6 DMRs in sex-stratified analysis. Shown are DMRs that are highly significant in one sex (FDR < 0.05), but not significant (P-value > 0.05) in another sex. Only 6 DMRs satisfied these criteria in male samples. For each DMR, annotations include location of the DMR based on hg19/GRCh37 genomic annotation (DMR), Illumina gene annotations, nearby genes based on GREAT, and chromatin state. The inverse-variance weighted meta-analysis regression models results based on coMethDMR include estimated effect size (estimate) where DMRs that are hyper-methylated in AD have positive values, its associated standard error (se), P-value, and false discovery rate (FDR) for multiple comparison corrections. Shown in bold are significant results with FDR < 0.05.

In sex-by-Braak stage interaction analysis, we identified significant interaction at 14 CpGs, but no significant interactions at DMRs at 5% FDR. There was also little overlap between significant DNA methylation changes identified in sex-stratified and sex-by-Braak stage interaction analyses. Only 4 CpGs were identified by both analyses (Table 3). To understand this discrepancy, note that the sex-stratified analysis detected many changes that are attenuated but might be in the same direction in one sex group compared to the other. For example, among the 10 most significant CpGs from the sex-stratified analysis, 9 female-specific and 6 male-specific CpGs (Table 1) had the same direction of methylation-Braak stage association in both sexes. On the other hand, in sex-by-Braak stage interaction analysis, 13 out of the 14 significant CpGs had the opposite directions of changes for methylation-Braak stage associations in females and males (Table 3). Therefore, the interaction analysis was able to identify CpGs (or DMRs) with large differences in sex-specific effect estimates, often in different directions, but these effects might not have reached FDR significance in sex-stratified analysis. For example, from Table 3, the CpG with the most significant interaction (cg13212831) had effect estimates 0.083 and −0.139 for females and males, respectively. In sex-stratified analysis, although the methylation-Braak stage associations were highly significant (P-value_female_ = 0.006, P-value_male_ = 4.1 × 10^−5^), they did not reach 5% FDR significance threshold (FDR_female_ = 0.413, FDR_male_ = 0.097). Therefore, the results from sex-stratified analysis and sex-by-Braak stage interaction analysis complemented each other.

**Table 3.** Results from sex-by-Braak stage interaction analysis. For each CpG, annotations include the location of the CpG based on hg19/GRCh37 genomic annotation (chr, position), Illumina gene annotations, nearby genes based on GREAT, and chromatin state. The inverse-variance weighted meta-analysis regression models results include estimated effect size (estimate) where CpGs that are hyper-methylated in AD have positive values, its associated standard error (se), and P-value. Adj.pval is adjusted P-value from stageR analysis. Shown in bold are significant results (adj.pval < 0.05 for interaction or P-value < 0.05 for one sex). * indicates these CpGs also reached 5% FDR significance in sex-stratified analysis.

| cpg | Annotations |  |  |  |  | Female samples |  |  | Male samples |  |  | Sex * Braak stage interaction |  |  |  |
| --- | --- | --- | --- | --- | --- | --- | --- | --- | --- | --- | --- | --- | --- | --- | --- |
|  | chr | Position | Illumina | GREAT (distance to TSS) | chromatin state | estimate | se | P-value | estimate | se | P-value | estimate | se | P-value | adj.pval |
| cg13212831 | chr1 | 56046343 |  | USP24(-365558); PPAP2B(+998897) | Weak transcription | 0.083 | 0.030 | **6.15E-03** | -0.139 | 0.034 | **4.10E-05** | -0.222 | 0.044 | **6.38E-07** | **3.54E-04** |
| cg25734825 | chr3 | 119182523 | TMEM39A | TMEM39A(+5) | Active TSS | 0.059 | 0.030 | 5.01E-02 | -0.156 | 0.036 | **1.21E-05** | -0.220 | 0.045 | **1.12E-06** | **6.22E-04** |
| *cg02331272 | chr11 | 66964208 | KDM2A;KDM2A | ADRBK1(-69672); KDM2A(+77051) | Weak transcription | 0.058 | 0.030 | 1.10E-01 | -0.166 | 0.036 | **2.99E-06** | -0.215 | 0.046 | **2.25E-06** | **1.25E-03** |
| cg10622825 | chr1 | 172413349 | PIGC;PIGC;C1orf10 | PIGC(-124) | Active TSS | -0.069 | 0.030 | **2.22E-02** | 0.134 | 0.034 | **7.78E-05** | 0.200 | 0.045 | **7.00E-06** | **3.88E-03** |
| cg10784067 | chr16 | 14014435 | ERCC4 | ERCC4(+422) | Active TSS | -0.127 | 0.032 | **5.33E-05** | 0.068 | 0.033 | **3.78E-02** | 0.201 | 0.045 | **9.03E-06** | **5.01E-03** |
| cg14284055 | chr1 | 24439399 | MYOM3 | MYOM3(-735) | Bivalent Enhancer | 0.115 | 0.029 | **8.28E-05** | -0.066 | 0.037 | 7.12E-02 | -0.193 | 0.045 | **1.94E-05** | **1.07E-02** |
| *cg18942110 | chr15 | 91072797 | CRTC3;CRTC3 | CRTC3(-502) | Active TSS | 0.031 | 0.030 | 3.06E-01 | -0.164 | 0.035 | **2.23E-06** | -0.187 | 0.045 | **3.13E-05** | **1.74E-02** |
| cg09342330 | chr14 | 89021109 | PTPN21;PTPN21 | PTPN21(-33) | Active TSS | 0.045 | 0.030 | 1.31E-01 | -0.144 | 0.037 | **8.30E-05** | -0.189 | 0.046 | **3.47E-05** | **1.93E-02** |
| cg21722170 | chr6 | 31977443 | TNXB;TNXA | C4B(-5095); C4A(+27643) | Quiescent/Low | 0.034 | 0.030 | 2.53E-01 | -0.148 | 0.034 | **1.57E-05** | -0.183 | 0.045 | **4.17E-05** | **2.31E-02** |
| cg03662217 | chr21 | 35747882 | FAM165B | SMIM11(-20) | Active TSS | 0.039 | 0.029 | 1.85E-01 | -0.141 | 0.036 | **7.27E-05** | -0.184 | 0.045 | **4.59E-05** | **2.55E-02** |
| cg15026265 | chr12 | 28283812 |  | PTHLH(-158910); CCDC91(-59566) | Quiescent/Low | -0.038 | 0.029 | 1.95E-01 | 0.134 | 0.033 | **6.58E-05** | 0.176 | 0.044 | **6.72E-05** | **3.73E-02** |
| cg00659421 | chr7 | 86781760 | DMTF1;DMTF1;DM | DMTF1(-109) | Active TSS | -0.050 | 0.031 | 1.08E-01 | 0.130 | 0.033 | **7.53E-05** | 0.178 | 0.045 | **6.95E-05** | **3.86E-02** |
| *cg24917065 | chr8 | 23418389 | SLC25A37 | SLC25A37(+32072); NKX3-1(+12205) | Quiescent/Low | -0.135 | 0.030 | **6.26E-06** | 0.042 | 0.033 | 2.05E-01 | 0.174 | 0.044 | **6.98E-05** | **3.87E-02** |
| *cg18281939 | chr5 | 77783895 | LHFPL2 | SCAMP1(+127557); LHFPL2(+16075) | Weak transcription | -0.022 | 0.029 | 4.55E-01 | -0.179 | 0.036 | **6.26E-07** | -0.177 | 0.045 | **7.10E-05** | **3.94E-02** |

Consistent with previous studies [18, 19, 36, 37], we observed the majority of these sex-specific changes were hyper-methylated in AD samples, for which methylation levels increased as AD Braak stage increased. More specifically, 59% of the significant CpGs and 69% of the significant DMRs in females, along with 66% of the significant CpGs and 89% of the significant DMRs in males were hyper-methylated in AD (Supplementary Table 3-6).

### Enrichment analysis of sex-specific DNA methylation changes in different genomic features

Compared to background probes, significant hypermethylated DMRs and CpGs in females are over-represented in CpG islands and gene bodies and under-represented in open sea and intergenic regions (Supplementary Figure 2a, Supplementary Table 7). In contrast, significant hypermethylated DMRs and CpGs in males are over-represented in shores, 5’UTRs and TSS1500s, but under-represented in open seas, gene bodies, and intergenic regions.

Significant hypomethylated changes in females are over-represented in open seas, but under-represented in CpG islands, TSS200s, and intergenic regions, while significant hypomethylated changes in males are over-represented in open seas (Supplementary Figure 2b, Supplementary Table 7). Interestingly, compared to males, significant hyper-methylated changes in females are more likely to occur in CpG islands and gene bodies and less likely to occur in shores and 5’UTRs. On the other hand, significant hypomethylated CpGs in females are less likely to occur in CpG islands.

Our enrichment analysis with respect to chromatin states showed that significant hyper-methylated changes in females were enriched in bivalent enhancer, flanking active TSS, repressed polycomb, and transcription at gene 5’ and 3’ regions, but depleted in quiescent/low, strong transcription, weakly repressed polycomb, and weak transcription regions (Supplementary Figure 2c, Supplementary Table 8). In contrast, significant hypermethylated changes in males were enriched in active TSS, flanking active TSS, and repressed polycomb regions, but depleted in quiescent/low, strong transcription, weak repressed polycomb, and weak transcription regions.

Hypomethylated changes in females were enriched in enhancers, weak repressed polycomb, and weak transcription regions, but depleted in active TSS, bivalent/poised TSS, and repressed polycomb regions, while hypomethylated changes in males were enriched in flanking active TSS and weak repressed polycomb regions (Supplementary Figure 2d, Supplementary Table 8). Compared to males, significant hyper-methylated changes in females are more likely to occur at bivalent enhancer and flaking active TSS, but less likely to occur in active TSS, flanking bivalent TSS/Enh, and quiescent/low regions, while significant hypomethylated changes in females are less likely to occur at active TSS and weakly repressed polycomb regions.

Similarly, enrichment tests for regulatory elements using the LOLA software also supported the potential functional relevance of these significant changes in DNA methylation. Significant DMRs and CpGs in females and males were both enriched in binding sites of EZH2 and SUZ12 (Supplementary Table 9), which are subunits of polycomb repressive complex 2 (PRC2), consistent with the observed enrichment of methylation differences in PRC2 repressed regions (Supplementary Figure 2c,d) and previous observations that DNA methylation often interact with PRC2 binding [38, 39]. Other top hits included binding sites for SPI1, TCF7L2, CEBPB, and CtBP2 in females, and MafK in males.

### Gene ontology and pathway analysis

Because of the relatively smaller number of gene sets being tested, a 25% FDR significance threshold, instead of the conventional 5% FDR, was suggested for GSEA [40]. At 25% FDR, the significant DNA methylation changes in females were enriched in `TYROBP causal network` (FDR = 0.014), `TCR signaling in naïve CD4+ T cells` (FDR = 0.130) and `ROBO receptors bind AKAP5` (FDR = 0.160) gene sets, and significant methylation changes in males were enriched in `initial triggering of complement` gene set (FDR = 0.245). The TYROBP causal network was previously inferred from a large-scale network analysis of human LOAD brains [41]; it was FDR significant (P-value < 0.001, FDR = 0.014) in females (Supplementary Figure 3a), compared to a nominal association in males (P-value < 0.001, FDR = 0.620). Interestingly, the core enrichment subset of genes identified by GSEA in the female and male subnetworks regulated by TYROBP involved DNA methylation changes at different genes (Supplementary Figure 3b), highlighting different regulatory mechanisms for this gene network in males and females.

The comparison with gene ontology (GO) terms showed at 25% FDR, significant methylation changes in females were enriched in 25 GO biological processes (Table 4, Supplementary Table 10), many of which are involved in inflammatory responses associated with AD pathology including `CD8 positive alpha beta T cell activation` and `interferon alpha production,` as well as other biological processes critical for AD pathogenesis such as `response to platelet derived growth factor` and `positive regulation of axon extension.` For males, we did not identify any significant GO terms at 25% FDR; the strongest enrichment with nominal P-value less than 0.001 involved immune responses to the accumulation of amyloid-β (Aβ) in the brain, such as `regulation of T cell activation via T cell receptor contact with antigen bound to MHC molecule on antigen presenting cell,` and other biological processes recently implicated in AD such as response to angiotensin [42, 43] and cell redox homeostasis [44, 45].

**Table 4.** Top 10 GO Biological processes enriched with sex-specific DNA methylation changes associated with AD Braak stage in females and males. Shown are GSEA results including number of genes in the gene set (SIZE), normalized enrichment score (NES), P-value, FDR and relevant AD literature for the gene set.

| Gene Set | SIZE | NES | P-value | FDR | Relevance to AD |
| --- | --- | --- | --- | --- | --- |
| **<u>Top 10 GO Biological Process terms in females</u>** |  |  |  |  |  |
| INTEGRIN_ACTIVATION | 24 | 2.105 | 0 | 9.53E-02 | Wennströme et al. (2012)[92] |
| RESPONSE_TO_PLATELET_DERIVED_GROWTH_FACTOR | 19 | 2.116 | 0 | 1.07E-01 | Sil et al. (2018)[93] |
| I_KAPPAB_PHOSPHORYLATION | 19 | 2.081 | 0 | 1.13E-01 | Jha et al. (2019)[94] |
| NEGATIVE_REGULATION_OF_INTERLEUKIN_8_PRODUCTION | 18 | 2.133 | 0 | 1.21E-01 | Qin et al. (2016)[95] |
| POSITIVE_REGULATION_OF_MACROPHAGE_MIGRATION | 25 | 2.048 | 0 | 1.46E-01 | Bacher et al. (2010) [96] |
| TOLL LIKE RECEPTOR SIGNALING PATHWAY | 142 | 2.031 | 0 | 1.61E-01 | Fiebich et al. (2018)[97] |
| NEGATIVE_REGULATION_OF_TUMOR_NECROSIS_FACTOR_SUPERFAMILY_CYTOKINE_PRODUCTION | 57 | 2.007 | 0 | 1.90E-01 | Chang et al. (2017) [98] |
| REGULATION_OF_SYNCYTIIUM_FORMATION_BY_PLASMA_MEMBRANE_FUSION | 28 | 1.944 | 0 | 1.98E-01 | Armoto et al. (2013)[99] |
| RESPONSE_TO_VITAMIN_A | 19 | 1.945 | 0 | 2.08E-01 | Ono et al. (2012)[100] |
| POSITIVE_REGULATION_OF_AXON_EXTENSION | 42 | 1.931 | 0 | 2.08E-01 | Kanaan et al. (2013)[101] |
| **<u>Top 10 Biological Process terms in males</u>** |  |  |  |  |  |
| REGULATION_OF_T_CELL_ACTIVATION_VIA_T_CELL_RECEPTOR_CONTACT_WITH_ANTIGEN_BOUND_TO_MHC_MOLECULE_ON_ANTIGEN_PRESENTING_CELL | 6 | 1.869 | 0 | 5.98E-01 | Schettters et al. (2017)[102] |
| REGULATION_OF_SYSTEMIC_ARTERIAL_BLOOD_PRESSURE_BY_CIRCULATORY_RENIN_ANGIOTENSIN | 18 | 1.964 | 0 | 6.11E-01 | Cosarderelioglu et al. (2020)[42] |
| NEGATIVE_REGULATION_OF_REACTIVE_OXYGEN_SPECIES_BIOSYNTHETIC_PROCESS | 29 | 1.745 | 0 | 6.12E-01 | Manoharan et al. (2016)[44] |
| COMPLEMENT_ACTIVATION | 68 | 1.741 | 0 | 6.26E-01 | Morgan (2018)[80] |
| RESPONSE_TO_ANGIOTENSIN | 26 | 1.746 | 0 | 6.36E-01 | Benigni et al. (2010)[43] |
| CELL_REDOX_HOMEOSTASIS | 55 | 1.721 | 0 | 6.49E-01 | Chen et al. (2020)[45] |
| PROTEIN_DEMETHYLATION | 28 | 1.832 | 0 | 6.77E-01 | Esposito et al. (2019)[103] |
| IMMUNE_RESPONSE_INHIBITING_CELL_SURFACE_RECEPTOR_SIGNALING_PATHWAY | 6 | 1.781 | 0 | 7.02E-01 | Schettters et al. (2017)[102] |
| DICARBOXYLIC_ACID_CATABOLIC_PROCESS | 17 | 2.052 | 0 | 7.57E-01 | Griffin et al. (2017)[104] |
| GLUTAMINE_FAMILY_AMINO_ACID_METABOLIC_PROCESS | 70 | 1.657 | 0 | 7.75E-01 | Conway et al. (2020)[105] |

### Correlation of sex-specific DNA methylation changes in AD with expression levels of nearby genes

Using the ROSMAP dataset with 529 samples (333 females and 196 males) with both DNA methylation and RNA-seq gene expression data, we next evaluated the role of significant DMRs or CpGs by correlating their DNA methylation levels to the expression of genes linked by GREAT [32], which associates genomic regions to target genes. At 5% FDR, for FDR significant CpGs in females, out of the 381 CpGs that were linked to a nearby gene, 14 were significantly associated with target gene expression levels (Supplementary Table 11), and half of them (n = 7) had effects in the negative direction. Among FDR significant CpGs in males, out of the 46 CpGs that were linked to a nearby gene, 2 were significantly associated with target gene expression and both were in the negative direction. Notably, in females, several of the most significant CpG methylation-gene expression associations were observed for the *HLA-DPA1* gene, which encodes microglia receptors involved in antigen presentation and is regulated by PU.1 [46]. In males, the most significant CpG-gene expression was for *HLA-DRB1*, another PU.1 target gene [46]. For the 14 CpGs identified by our sex-by-Braak stage interaction analysis, only one CpG (cg24917065) was significantly associated with target gene (*SLC25A37*) expressions.

### Correlation and overlap of sex-specific DNA methylation changes in AD with genetic susceptibility loci

To evaluate if the significant methylation differences are located in the vicinity of sex-specific genetic variants implicated in AD, we compared our sex-specific CpGs and DMRs with the recently identified sex-specific SNPs associated with AD biomarkers [47] or AD pathology [48]. We found only 5 CpGs, mapped to the *SERP2, KCNE1, TNKS1BP1, FAM165B, PLCB4* genes were located within 500 kb of the sex-specific SNPs (Supplementary Table 12).

To search for mQTLs, we next tested associations between the sex-specific CpGs and DMRs with SNPs that are located within 500 kb from them using 688 samples (434 females, 254 males) from the ROSMAP study, which had both genotype and DNA methylation data. While no mQTL-DMR pairs reached 5% FDR significance, we did identify 572 and 284 FDR-significant mQTL-CpG pairs associated with the sex-specific CpGs in females and males, respectively (Supplementary Tables 13-15). Among the 381 and 76 sex-specific CpGs identified in female and male samples, respectively, 41 (11%) and 15 (20%) had at least one corresponding mQTL in brain samples. Among the 14 CpGs identified in our sex-by-Braak stage interaction analysis, 2 and 7 CpGs with at least one brain mQTL, corresponding to 21 and 236 significant mQTL-CpG pairs, were identified at 5% FDR in females and males, respectively.

### Drug target analysis of sex-specific DNA methylation changes

To investigate the clinical impact of the sex-specific DNA methylation changes, we next compared them with targets of drugs in the ChEMBL database [34] that are annotated to Alzheimer disease, many of which are antipsychotic medications commonly prescribed to AD patients for treating psychiatric symptoms that accompany AD. We found that 13 CpGs and 2 DMRs, mapped to 20 genes, had overlap with targets of 16 different drugs (Supplementary Table 16). Among them, *CACNA1C* encodes voltage-dependent calcium channel, which is a target of cholinesterase inhibitor donepezil. Previously, drug response for donepezil were shown to be modulated by the sex hormone estrogen receptor alpha (ESR1) genotype [49]. Also, *CHRM3* encodes muscarinic acetylcholine receptor, which is targeted by two commonly prescribed antipsychotic drugs for AD patients, trazodone and haloperidol. In both human and animal models, it has been observed treatment with haloperidol induces sex-specific DNA methylation changes [50, 51]. Several CpGs and one DMR are mapped to targets of valproic acid, a mood stabilizer often prescribed for AD patients and was shown to have different pharmacokinetic profiles between male and female subjects [52]. Interestingly, two CpGs and 1 DMR also mapped to targets of caffeine, which was included in cocktail therapy in AD clinical trials [53, 54]. Although caffeine reduces the risk for AD [55, 56] in both men and women, the protective effect seem to be greater in women [57]. Taken together, these results highlighted the clinical importance of the sex-specific DNA methylation changes.

## Discussion

To identify sex-specific changes in AD, we employed two complementary approaches, a sex-stratified analysis that examined methylation-Braak stage associations in female and male samples separately, and a sex-by-Braak stage interaction analysis that compared the magnitude of these associations between different sexes. In sex-stratified analysis, as discussed above, a substantial number of the significant loci showed the same direction but attenuation of effect size for methylation-Braak stage association in a different sex (Table 1 and 2). Therefore, it is not surprising that many of these significant CpGs were identified previously in sex-combined meta-analysis [22]. Among FDR significant methylation changes in females, 325 CpGs (85%) and 40 DMRs (56%), mapped to genes such as *HOXA3, AZU1,* and *MBP,* were also previously identified in our sex-combined meta-analysis [22] (Supplementary Tables 3 and 5). Similarly, in the analysis of male samples, among FDR significant changes, 58 (76%) CpGs and 15 DMRs (56%), mapped to genes such as *MAMSTR, RHBDF2,* and *AGAP2,* overlapped with significant hits from sex-combined meta-analysis [22] (Supplementary Tables 4 and 6). However, our sex-specific analysis provided the new insight that the effects of these known AD genes appear to be predominately driven by effects in only one sex (Table 1 and 2).

On the other hand, our sex-specific analysis also uncovered novel methylation changes at 84 CpGs and 42 DMRs that were not identified previously by sex-combined analyses[22], which might have reduced power due to heterogeneity between the sexes. For example, among the top 10 CpGs in the sex-stratified analysis (Table 1), a new locus at cg22632947, which mapped to the gene body of the *PRKCA* gene, was highly significant in female samples (estimate = −0.139, P-value = 1.50 × 10^−7^, FDR = 3.00 × 10^−3^), but not significant in male samples (estimate = −0.005, P-value = 0.857, FDR = 0.995) (Supplementary Figure 4). The *PRKCA* gene encodes protein kinase Cα (PKCα), which participates in synaptic loss resulting from accumulation of amyloid-β (Aβ) in AD [58, 59]. Another novel locus is at cg18942110 in the promoter of the *CRTC3* gene, where methylation-Braak stage association was highly significant in male samples (estimate = −0.164, P-value = 2.23 × 10^−6^, FDR = 3.19 × 10^−2^), but not significant in female samples (estimate = −0.031, P-value = 0.306, FDR = 0.952) (Supplementary Figure 4). CRTC3 is a member of the CRTC family, which are coactivators of the transcription factor CREB (cAMP-response element binding protein). In addition to its crucial role in maintaining synaptic plasticity and facilitation of short-term memory to long term memory, the CREB signaling pathway also mediates synapse loss induced by Aβ in AD [60]. Notably, synapse loss significantly correlates with cognitive impairment [61, 62] and has been observed to be an early feature of AD pathogenesis [63, 64].

The sex-by-Braak stage interaction analysis also uncovered a number of additional novel methylation loci that affected AD in a sex-specific manner. Notably, none of the 14 CpGs detected in our interaction analysis were identified in previous large-scale DNA methylation studies [18–22], suggesting that sex-specific changes such as these can be missed by conventional studies that do not consider the impact of sex. This is likely due to cancelation of effects in sex-combined analysis, because the majority of these 14 CpGs had different directions of methylation-Braak stage effects in male and female samples (Table 3). Among genes mapped to these 14 CpGs, TMEM39A is a member of the transmembrane (TMEM) protein family. In recent GWAS, a genetic variant on *TMEM39A* was discovered and replicated as an important risk locus for multiple sclerosis, an autoimmune condition of the central nervous system [65, 66]. While relatively little is known about the role of *TMEM39A* in AD, given its important contributions to inflammation, dysregulated type I interferon responses, and other immune processes [67] which are also implicated in AD, methylation differences affecting this gene are particularly relevant. Another noteworthy gene is *TNXB* and its pseudo gene *TNXA*, which are located in the major histocompatibility complex (MHC) class III region on chromosome 6. *TNXB* encodes tenascin proteins, which are extracellular matrix glycoproteins demonstrated to modulate synaptic plasticity in the brain [68]. In particular, genetic variants at the *HLA-DQB1* locus discovered in the recent AD genetic meta-analysis [69] included eQTLs for *TNXB/TNXA* in brain tissues [69, 70].

To better understand the relevance of these AD-associated sex-specific changes, we also compared our results with several previous studies. The comparison with Xia et al. (2019) [16] and Xu et al. (2014) [17], which examined differential methylation between males and females in the prefrontal cortex, but without considering AD status [16, 17], showed our results were largely distinct. Among 451 unique CpGs identified in our sex-stratified analysis or sex-by-Braak stage interaction analysis, only 16 were also identified in Xia et al. (2019) [16] and none were identified in Xu et al. (2014) [17] (Supplementary Tables 3-6). This is probably due to different hypotheses tested in our study and the sexual dimorphism studies – while our study examined the impact of sex on methylation-Braak stage association, the previous studies examined differential methylation between the sexes, regardless of AD severity. The comparison of our results with sex-specific DNA methylation changes in fetal brain development [71, 72] also showed very little overlap (Supplementary Table 17); one hypothesis could be that the AD-associated sex-specific DNA methylation changes identified in this study might be influenced by environmental risk factors for AD, such as diet and exercise.

The results of our gene set analysis highlighted a number of critical sex-specific biological processes in AD. Notably, the TYROBP causal network reached FDR significance threshold in females (FDR = 0.014) but was only nominally significant in males. Interestingly, AD-associated CpG methylation changes that drove pathway associations (core enrichment genes) occurred at different genes in females and males (Supplementary Figure 3), indicating a potentially sex-specific regulatory mechanism for this network. TYROBP (TYRO protein tyrosine kinase-binding protein) is a key regulator of the complement pathway in the immune/microglia network, which is activated as Aβ accumulates in LOAD brains [41, 73]. TYROBP is a transmembrane adaptor protein for TREM2, SIRPβ1, and CR3 receptors, which are known to be involved in AD pathogenesis [73–75]. In addition, TYROBP is regulated by SPI1, a central hub for the network of genes involved in myeloid immune response in neurodegeneration [76]. In patients with LOAD, TYROBP was observed to be up-regulated in the brains in multiple cohorts [41]. Recent studies suggested TYROBP-mediated signaling is involved in multiple important functions as aggregating Aβ activates microglia, including enhanced phagocytosis of damaged neurons [41, 73] and suppression of inflammatory responses [77], as well as neuronal pruning activity [41]. Interestingly, in gene ontology (GO) analysis, among the most significant GO Biological Process terms (P-value < 0.001) in females and males, none of them overlapped (Supplementary Table 10), even though the relevancy of all the top biological processes were supported by recent AD literature (Table 4). These results suggest different biological processes are associated with AD pathology in males and females.

Importantly, a number of these sex-specific biological processes pointed to important potential biomarkers and therapeutic targets for the treatment of AD. For example, one of the top biological process enriched with significant methylation changes in female samples is `response to platelet derived growth factor`. Recently, multiple studies have shown that reduced levels of platelet-derived growth factors (PDGFs) in plasma significantly correlate with mild cognitive impairment and so have proposed PDGFs as a potential biomarker for AD [78, 79]. For the significant methylation changes in male samples, one of the top biological process highlighted by our enrichment analysis is `dysregulation in the complement system`. Recently, a number of novel agents targeting the complement system are being developed and tested in clinical trials for potential effective therapy for AD [80]. Therefore, clinical trials testing potential treatment for AD patients might have more power for detecting treatment effects by considering sex and targeting the subgroup with the higher predicted benefit based on patient molecular profiles such as DNA methylation.

The comparison with ChEMBL database [34] showed a number of sex-specific DNA methylation changes were also located in regulatory regions of genes targeted by drugs often prescribed to AD patients, such as those treating neuropsychiatric symptoms accompanying AD. Several antipsychotic drugs, such as haloperidol, donepezil and valproic acid had targets in genomic regions where DNA methylation levels associated with AD in a sex-specific manner. Taken together, these results highlighted the importance of developing and applying sex-specific treatment regimens in AD.

Our study also provided support for the sex-specific effect of brain estrogen in AD. Among FDR significant CpGs in males is cg15626350, located on the gene body of *ESR1* gene, which encodes estrogen receptor alpha, one of two subtypes of estrogen receptor. Genetic polymorphisms of *ESR1* have been associated with risk of developing cognitive impairment in elders [81–85]. In addition, multiple animal and in vitro studies have demonstrated the neuroprotective effect of estrogen [86, 87], which promotes neurogenesis, neuronal plasticity, synaptic transmission and reduces Aβ production. Interestingly, cg15626350 reached 5% FDR significance threshold in males (estimate = 0.144, P-value = 2.54 × 10^−6^, FDR = 0.033), but is only nominally significant in females (estimate = 0.107, P-value = 5.08 × 10^−4^, FDR = 0.164), indicating a stronger methylation-AD Braak stage association in males (Supplementary Figure 4). Although not statistically significant, cg15626350 also showed stronger association with *ESR1* gene expression in males (estimate = 0.152, P-value = 0.067) compared to females (estimate = 0.031, P-value = 0.715). Importantly, the estrogen receptor is targeted by a number of drugs commonly prescribed for AD patients, such as trazodone, haloperidol, valproic acid (Supplementary Table 16). Together with the documented greater prevalence, disease severity and worse outcomes in females to infections and inflammation, particularly in the presence of reduced estradiol levels [88, 89], our findings suggest an interaction between estrogen levels and microglia activities, which may have led to altered inflammatory responses, ultimately resulting in sex differences in vulnerabilities to neurodegeneration in later life stages [90, 91]. Future experimental studies will help clarify the sex-specific epigenetic mechanisms of the estrogen pathway in modulating AD risk and drug responses.

There are several limitations for this study. The methylation levels in the studies analyzed here were measured on the bulk prefrontal cortex, which contains a complex mixture of cell types. To reduce confounding effects due to different cell types, we included estimated neuron proportion of each brain sample as a covariate variable in all our analyses. Currently, a challenge with cell-type specific studies is that they are often limited to smaller sample sizes due to labor-intensive sample preparation procedures and therefore have limited statistical power. Also, we did not identify any CpGs or DMRs from chromosome X, this might suggest that sex-differences in AD are not primarily due to chromosome X. Alternatively, the lack of association might also be due to the limited coverage by the 450k array. Future studies utilizing high throughput sequencing that provides better coverage of the epigenome will help clarify the role of the X chromosome in AD.

In summary, our study highlights the importance of stratifying on sex and analyzing sex-by-disease interaction in the analysis of DNA methylation data to discover the epigenetic architectures underlying AD. Our meta-analysis discovered a number of novel sex-specific DNA methylation changes consistently associated with AD Braak stage in multiple studies. Because of cancelation of effects in different directions, or dilution from samples with no effect, these sex-specific effects would be missed by sex-combined analysis. Moreover, for many genes previously linked to AD, our work provided evidence that the DNA methylation changes at these genes were predominately driven by effects in only one sex. Our enrichment analysis highlighted divergent biological processes in males and females, which underscored sex-specific regulatory mechanisms involved in AD. Finally, our results also have important implications for precision medicine - many of the sex-specific DNA methylation changes also pointed to important potential AD biomarkers and therapeutic targets, suggesting a pressing need for developing and applying sex-specific treatment strategies for AD.

## Data Availability

All datasets analyzed in this study are publicly available as described in Supplementary Table 1. The Mt. Sinai, London, Gasparoni and ROSMAP datasets can be accessed from GEO (accessions GSE80970, GSE59685, GSE66351) and Synapse (DOI:10.7303/syn3157275). The scripts for the analysis performed in this study can be accessed at https://github.com/TransBioInfoLab/ad-meta-analysis-by-sex

## Contributions

L.W., J.Y., E.R.M., L.Z., T.C.S., L.G. designed the computational analysis. L.Z., T.C.S., L.G., M.S., J.C., L.W. analyzed the data. L.W., J.Y., E.R.M, X.C. contributed to interpretation of the results. L.Z, L.W. wrote the paper, and all authors read and approved the manuscript. L.W. conceived the original idea and supervised the project.

## Ethics declarations

### Availability of data and materials

All datasets analyzed in this study are publicly available as described in Supplementary Table 1. The Mt. Sinai, London, Gasparoni and ROSMAP datasets can be accessed from GEO (accessions <u>GSE80970</u>, <u>GSE59685</u>, <u>GSE66351</u>) and Synapse (<u>DOI:10.7303/syn3157275</u>). The scripts for the analysis performed in this study can be accessed at https://github.com/TransBioInfoLab/ad-meta-analysis-by-sex.

### Ethics approval and consent to participate

Approval for ROSMAP dataset was obtained through Synapse. The ROSMAP study data were provided by the Rush Alzheimer’s Disease Center, Rush University Medical Center, Chicago. Data collection was supported through funding by NIA grants P30AG10161, R01AG15819, R01AG17917, R01AG30146, R01AG36836, U01AG32984, U01AG46152, the Illinois Department of Public Health, and the Translational Genomics Research Institute. The original Religious Orders Study and Rush Memory and Aging Project were approved by Institutional Review Board (IRB) of Rush University Medical Center.

### Consent for publication

Not applicable.

## Supplementary Text on Detailed Methods

### 1. Pre-processing of DNA methylation data

Quality control for CpG probes included removing probes with detection P-value < 0.01 in all samples of a cohort, those associated with cigarette smoking^1^, or those having a single nucleotide polymorphism (SNP) with minor allele frequency (MAF) 2: 0.01 present in the last 5 base pairs of the probe. Quality control for samples included restricting our analysis to samples with good bisulfite conversion efficiency (i.e., ≥ 88%) and principal component analysis (PCA). More specifically, PCA was performed using the 50,000 most variable CpGs for each cohort. Samples that were within ± 3 standard deviations from the mean of PC1 and PC2 were selected to be included in the final sample set. The quality controlled methylation datasets were next subjected to the QN.BMIQ normalization procedure^2^ as previously described^3^. The recorded sex status of all samples matched those predicted based on methylation levels using getSex function in R package minfi. We removed batch effects by applying linear model `methylation M value ∼ methylation slide.` For the ROSMAP cohort, we additionally included the variable “batch” that was available in the dataset to adjust for technical batches which occurred during data generation. The residuals (`methylation residuals)` estimated from this model were then used for subsequent analyses.

### 2. Single cohort analysis

To identify sex-specific DNA methylation changes in AD, we performed both a sex-stratified analysis and a sex-by-Braak stage interaction analysis for each of the four brain sample cohorts. In sex-stratified analysis, we tested methylation-Braak stage associations in female and male samples separately. In sex-by-Braak stage interaction analysis, we analyzed both female and male samples simultaneously and compared slopes for methylation-Braak stage associations in females and males.

In sex-stratified analysis, for each CpG, we applied the model methylation residuals ∼ age at death + Braak stage + CETS^4^ estimated neuron proportions to female samples and male samples separately. In sex-by-Braak stage interaction analysis, for each CpG, we applied the model `methylation residuals ∼ age at death + sex + Braak stage + sex*Braak stage + sex*age at death + CETS estimated neuron proportions` to samples of both sexes.

For the analysis of differentially methylated regions (DMRs), we used the coMethDMR R package^5^ to analyze 40,010 pre-defined genomic regions on the Illumina 450k arrays and identify co-methylated DMRs associated with Braak stage. The pre-defined genomic regions are regions on the Illumina array covered with clusters of contiguous CpGs where the maximum separation between any two consecutive probes is 200 base pairs. First, coMethDMR selects co-methylated sub-regions within these pre-defined contiguous genomic regions. Next, we summarized methylation M values within these co-methylated sub-regions using medians and tested them against AD Braak stage. The same linear models described for the analysis of CpGs were then applied to median value of each DMR.

### 3. Inflation assessment and correction

To assess inflation of the test statistics, we used quantile-quantile (QQ) plots of observed and expected distributions of P-values for each cohort. Because the conventional genomic inflation factor (lambda or λ used interchangeably below) is dependent on the expected number of true associations, and in a typical EWAS it is expected that small effects from many CpGs might be associated with the phenotype, Iterson et al. (2017)^6^ showed that the conventional genomic inflation factor would overestimate actual test-statistic inflation in EWAS. To estimate genomic inflations more accurately in EWAS, Iterson et al. (2017)^6^ developed a Bayesian method that estimates and corrects inflation in EWAS based on empirical null distributions, which is implemented in the Bioconductor package bacon. We estimated genomic inflation factors using both the conventional approach and the *bacon* method. In addition, we also applied the *bacon* method to single cohort analysis results to obtain inflation-corrected effect sizes, standard errors, and p-values for each cohort.

### 4. Meta-analysis

The results of the *bacon-*corrected cohort-specific analysis were then combined using inverse-variance weighted meta-analysis models. The evidence for heterogeneity of study effects was tested using Cochran’s Q statistic^7^. More specifically, the inverse-variance weighted fixed effects model was first applied to synthesize statistical significance from individual cohorts. Even though the fixed effects model for meta-analysis does not require the assumption of homogeneity^8^, for the regions with nominal evidence for heterogeneity (nominal P_heterogeneity_ < 0.05), we also applied random effects meta-analysis^9^ and assigned final meta-analysis P-value based on the random effects model. For each CpG (and for each DMR), we used the R package meta to obtain meta-analysis p-values for sex-by-Braak stage interaction, as well as Braak stage effect in female samples and male samples separately in sex-stratified analysis.

### 5. Identifying sex-specific changes

In sex-stratified analysis, we selected significant CpGs (or DMRs) with FDR < 0.05 in female samples or male samples separately. In sex-by-Braak stage interaction analysis, because the standard error of interaction effect `sex × Braak stage` is typically much larger than those for main `Braak stage` effects, the conventional approach for controlling false discovery rate often results in low power for discovering interaction effects^10^. Therefore, we used a stagewise analysis approach, previously proposed by van de Berge et al. (2017)^10^, to help improve power in high-throughput experiments where multiple hypotheses are tested for each gene. More specifically, in the *screening step*, for each CpG (or DMR), we tested the global null hypothesis that there is methylation-Braak stage association in either male or female samples. Next, in the *confirmation step*, we considered three individual null hypotheses for each CpG (or DMR): (a) there is no methylation-Braak stage association in male samples; (b) there is no methylation-Braak stage association in female samples; and (c) the methylation-Braak stage associations in male samples and female samples are the same. For the CpGs (or DMRs) selected in the screening step, these three individual hypotheses were then tested while controlling family-wise error rate (FWER) as described in van de Berge et al. (2017)^10^.

The stagewise analysis described above was implemented using the stageR package to identify CpGs (or DMRs) with significant differential methylation - Braak stage associations in females and males. In the screening step, we considered meta-analysis p-values for Braak stage in female samples and male samples (`p.meta.female`, `p.meta.male`), and used the minimum of these two meta-analysis p-values to represent each CpG (or DMR). In the confirmation step, the parameter pConfirmation was defined using three p-values for each CpG (or DMR): `p.meta.female`, `p.meta.male`, and `p.meta.interaction` (meta-analysis p-value for `sex × Braak stage`).

### 6. Enrichment and pathway analysis

The probes on the Illumina 450k array are annotated according to their locations with respect to genes (TSS1500, TSS200, 5’UTR, 1stExon, gene body, 3’UTR, intergenic) or to CpG islands (island, shore, shelf, open sea). To understand the genomic context of sex-specific DNA methylation changes in AD, we compared the FDR significant methylation changes from sex-stratified analysis with different types of genomic features. As pathological AD-associated methylation changes can occur at both significant individual CpGs and significant DMRs, we considered the CpGs located at significant individual CpGs or within significant DMRs jointly in this analysis, by testing their over- and under-representation in different types of genomic features using Fisher’s exact test. More specifically, the proportion of significant CpGs mapped to a particular type of genomic feature (e.g., CpG islands) (foreground) was compared to the proportion of CpGs on the array that mapped to the same type of genomic feature (background).

In addition, we used Fisher’s test to assess enrichment of significant CpGs and DMRs in different chromatin states by comparing with the 15-chromatin state data for DLPFC tissue samples (E073) from the Roadmap Epigenomics Project^11^. Using combinations of histone modification marks, ChromHMM^12^ was previously used to annotate segments of the genome with different chromatin states (repressed, poised and active promoters, strong and weak enhancers, putative insulators, transcribed regions, and large-scale repressed and inactive domains), which were shown to vary across sex, tissue type, and developmental age^13^. Similarly, we tested enrichment of significant CpGs and DMRs in binding sites of transcription factors and chromatin proteins from the ENCODE project^14^ and CODEX database^15^ using the LOLA R package^16^.

Finally, we performed pathway analysis by comparing the genes with significant DNA methylation changes in AD (identified in sex-stratified analysis) with the canonical pathways and biological process GO terms in MSigDB using GSEA analysis^17^. First, we linked each CpG and each pre-defined genomic region tested in DMR analysis (see Section “2. Single cohort analysis” above) to genes by annotating them using the GREAT (Genomic Regions Enrichment of Annotations Tool) software^18^ (with default “Basal plus method”), which associates genomic regions to target genes. Next, we represented each gene by the smallest p-value if multiple CpGs or genomic regions are associated with them. To remove selection bias due to different numbers of CpGs or genomic regions associated with each gene (i.e., P-values from a gene with many CpGs or genomic regions linked to it are likely to be smaller than a gene with few linked CpGs or DMRs), we next fit a generalized additive model^19^ using the R package mgcv: *Y_i_*∼*f*(*n. links_i_*) where *Y_i_* is negative log (base 10) transformation of the P-value for gene *i* in the analysis of female samples (or male samples), *n.links_i_* is the number of CpGs or DMRs associated with gene *i,* and *f* is a penalized spline function. We assumed gamma distribution for *Y_i_*, as under the null hypothesis of no association, *Y_i_* follows the chi-square distribution (a special case of gamma distribution). The residuals from this model were estimated and used to generate a ranked gene list, which was then used as input for GSEA (in pre-ranked mode) to identify canonical pathways and gene ontology terms (MsigDB C2:CP and C5:BP collections of gene sets) enriched with significant methylation changes in female samples and male samples separately.

### 7. Integrative methylation – gene expression analysis

To systematically evaluate transcriptional changes near the sex-specific DNA methylation changes, we next performed integrative methylation – gene expression analysis using 529 (333 female and 196 male) samples from the ROSMAP study with matched DNA methylation and gene expression data. To this end, normalized FPKM (Fragments Per Kilobase of transcript per Million mapped reads) gene expression values for the ROSMAP study were downloaded from the AMP-AD Knowledge Portal (Synapse ID: syn3388564). First, we linked significant CpGs (or DMRs) to nearby genes using GREAT^18^, which associates genomic regions to regulatory domains of genes. Next, we removed confounding effects in DNA methylation data by fitting the model `methylation M value ∼ neuron.proportion + batch + sample.plate + ageAtDeath` and extracting residuals from this model; these are the *methylation residuals*. Similarly, we also removed potential confounding effects in RNA-seq data by fitting model `log2(normalized FPKM values + 1) ∼ ageAtDeath + markers for cell types`. The last term, “markers for cell types,” included multiple covariate variables to adjust for the multiple types of cells in the brain samples. More specifically, we estimated expression levels of genes that are specific for the five main cell types present in the CNS: ENO2 for neurons, GFAP for astrocytes, CD68 for microglia, OLIG2 for oligodendrocytes, and CD34 for endothelial cells, and included these as variables in the above linear regression model, as was done in a previous large study of AD samples^20^. The residuals extracted from this model are the *gene expression residuals*.

Finally, for each gene expression and CpG (or DMR) pair, we then tested the association between gene expression residuals and methylation residuals using a linear model: `gene expression residuals ∼ methylation residuals + Braak stage`. For significant DMRs, this analysis was repeated, except that `methylation M value` was replaced with median methylation M value from multiple CpGs in the DMR.

### 8. Sex-specific mQTL analysis

To identify methylation quantitative trait loci (mQTLs) for the significant DMRs and CpGs, we tested associations between the methylation levels with nearby SNPs, using the ROSMAP study dataset, which had matched genotype data and DNA methylation data for 688 samples (434 females, 254 males). ROSMAP genotype data was downloaded from AMP-AD (syn3157325) and imputed to the Haplotype Reference Consortium r1.1 reference panel^21^. The male samples and female samples were analyzed separately.

To reduce the number of tests, we focused on identifying *cis* mQTLs located within 500kb from the start or end of the DMR (or position of the significant CpG)^22^. We additionally required SNPs to (1) have minor allele frequency of at least 1%, (2) be imputed with good certainty: information metric (info score) 2: 0.4, and (3) be associated with AD case-control status (as determined by clinical consensus diagnosis of cognitive status), after adjusting for age, batch, and the first 3 PCs estimated from genotype data, at nominal P-value less than 0.05. We then fit the linear model `methylation residual ∼ SNP dosage + batch + PC1 + PC2 + PC3`, where PC1, PC2, and PC3 are the first three PCs estimated from genotype data, to test the association between methylation residuals in CpGs and the imputed allele dosages for SNPs to identify mQTLs. The analysis for DMRs was the same except that we replaced methylation residual with median (methylation residuals) of all CpGs located within the DMR.

---

**Supplementary Figure 1.**
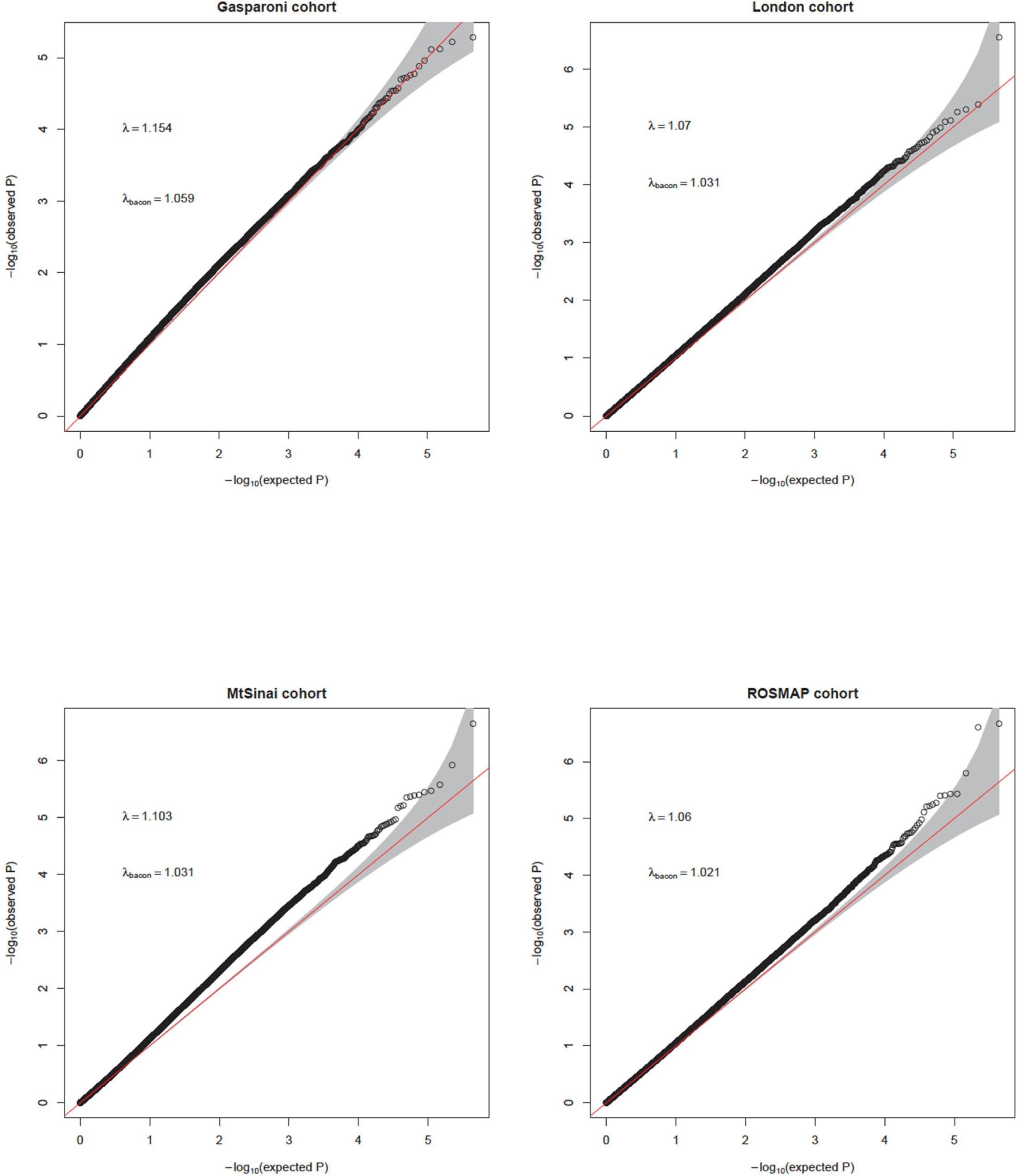
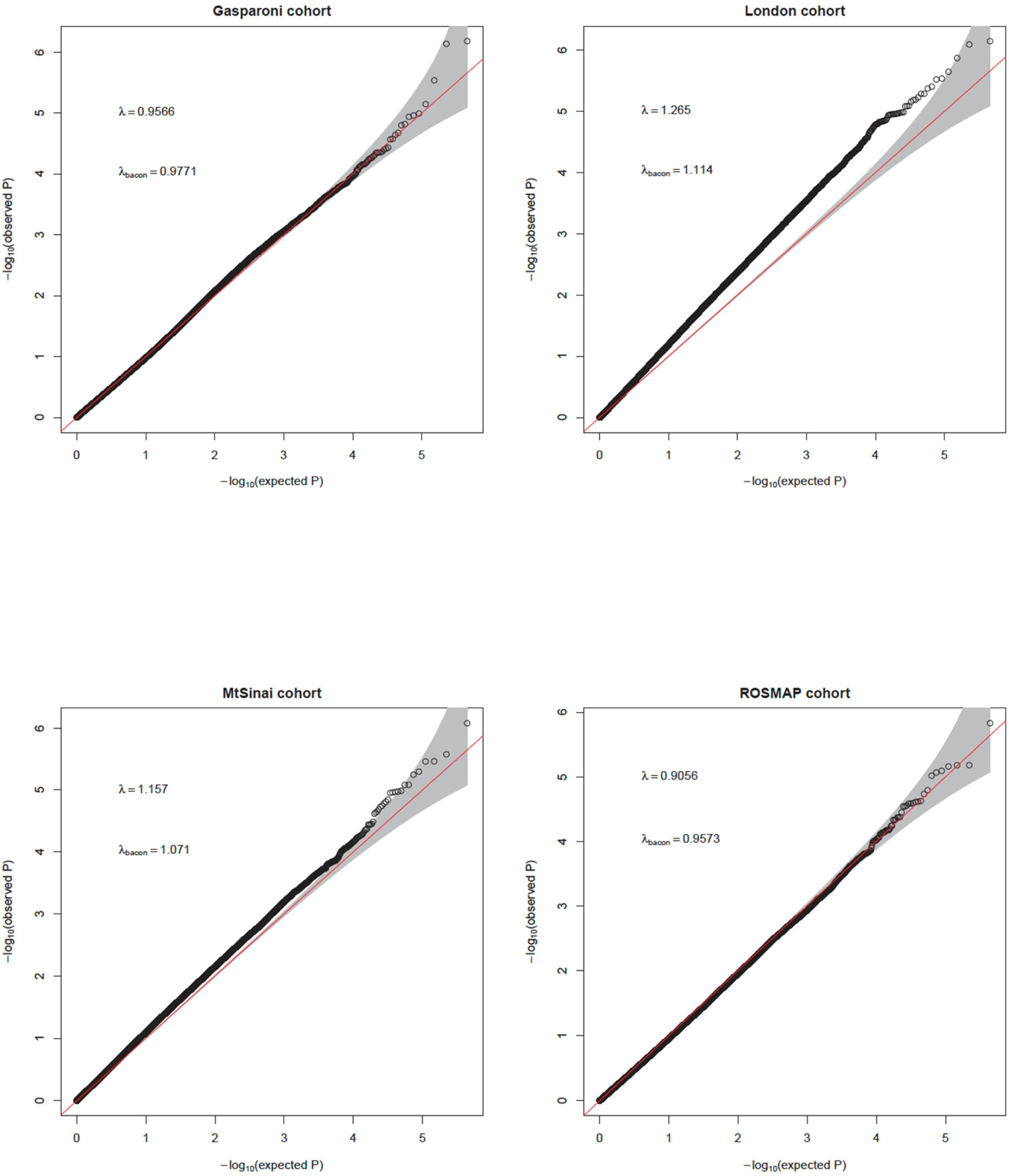
Quantile-quantile (QQ) plots of observed and expected distributions of p-values in Gasparoni, London, Mount Sinai, and ROSMAP cohorts. λ is the genomic inflation factor, and λ_bacon_ is the genomic inflation factor estimated using the method of Iterson et al. (2017) (PMID: 28129774), as implemented in the bacon R package. Shading indicates 95% confidence intervals. Reference line in red indicates expected distribution of −log_10_(P-values) under the null hypothesis of no association. (A) for analysis results of female samples (B) for analysis results of male samples

**Supplementary Table 2.**
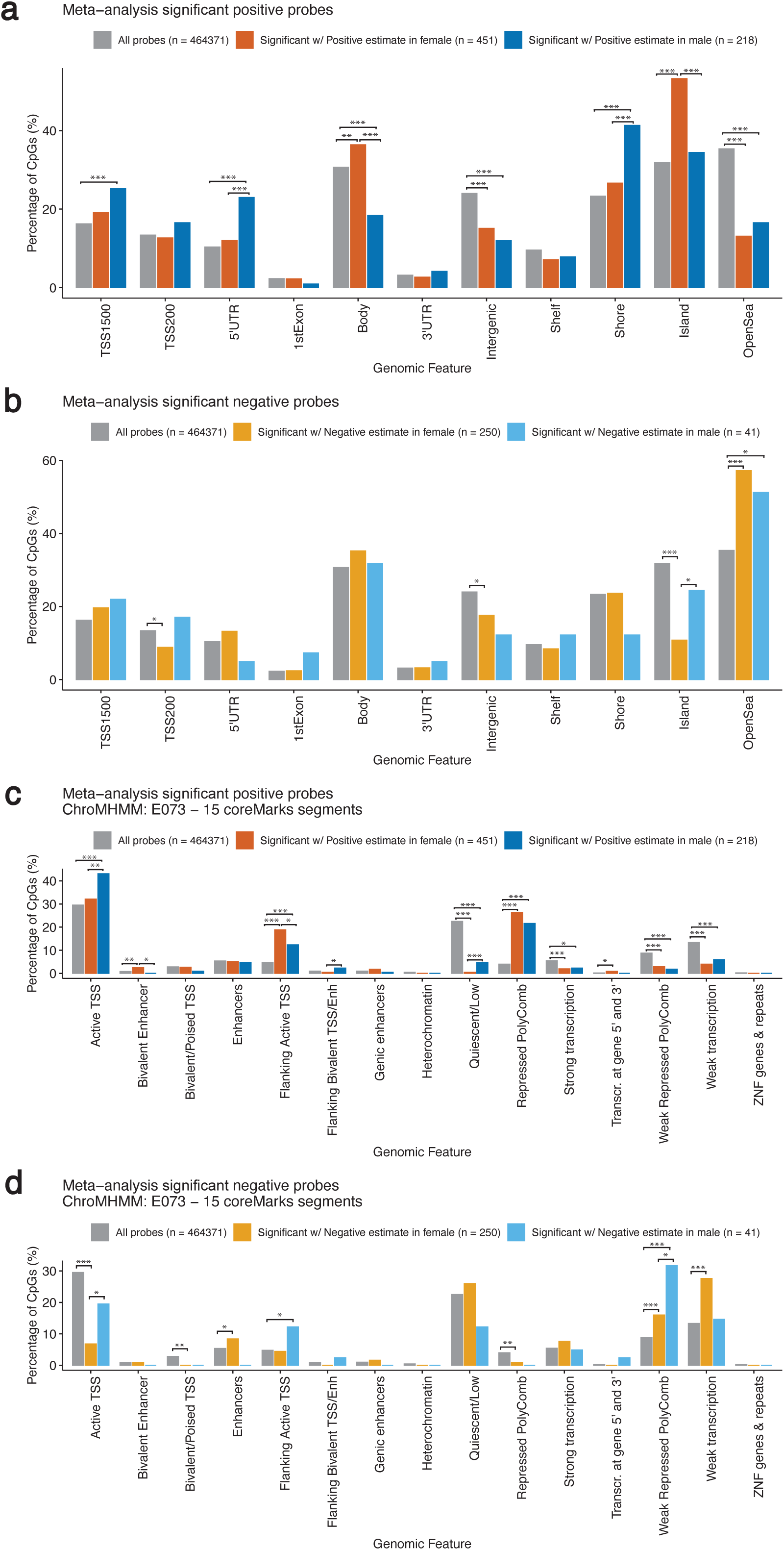
Enrichment of FDR significant CpGs and CpGs located within FDR significant DMRs with positive and negative effect estimates in various (A) (B) genomic features and (C) (D) chromatin states. *** indicates P-value < 0.001, ** indicates P-value < 0.01, * indicates P-value < 0.05

**Supplementary Figure 3.**
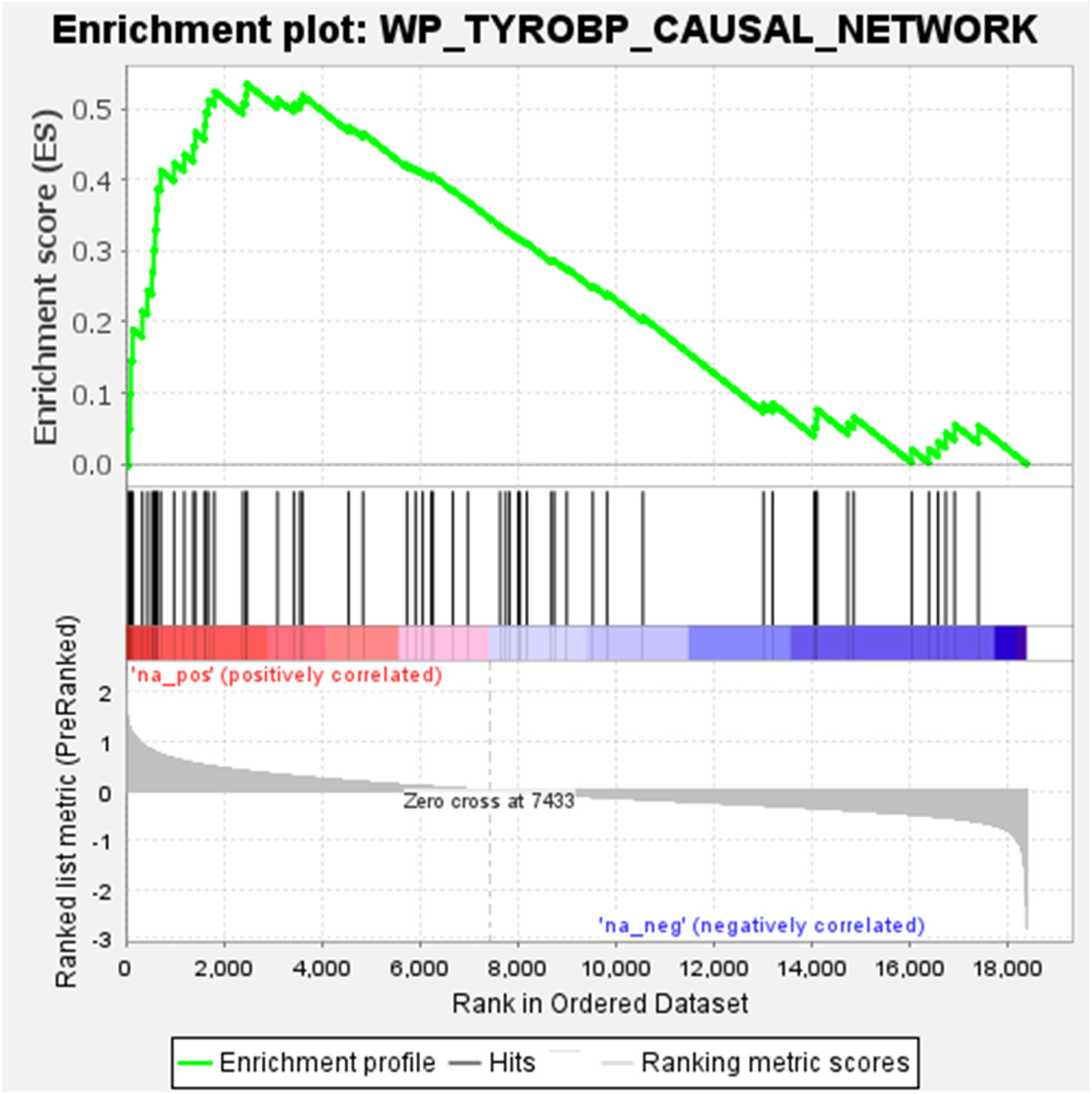
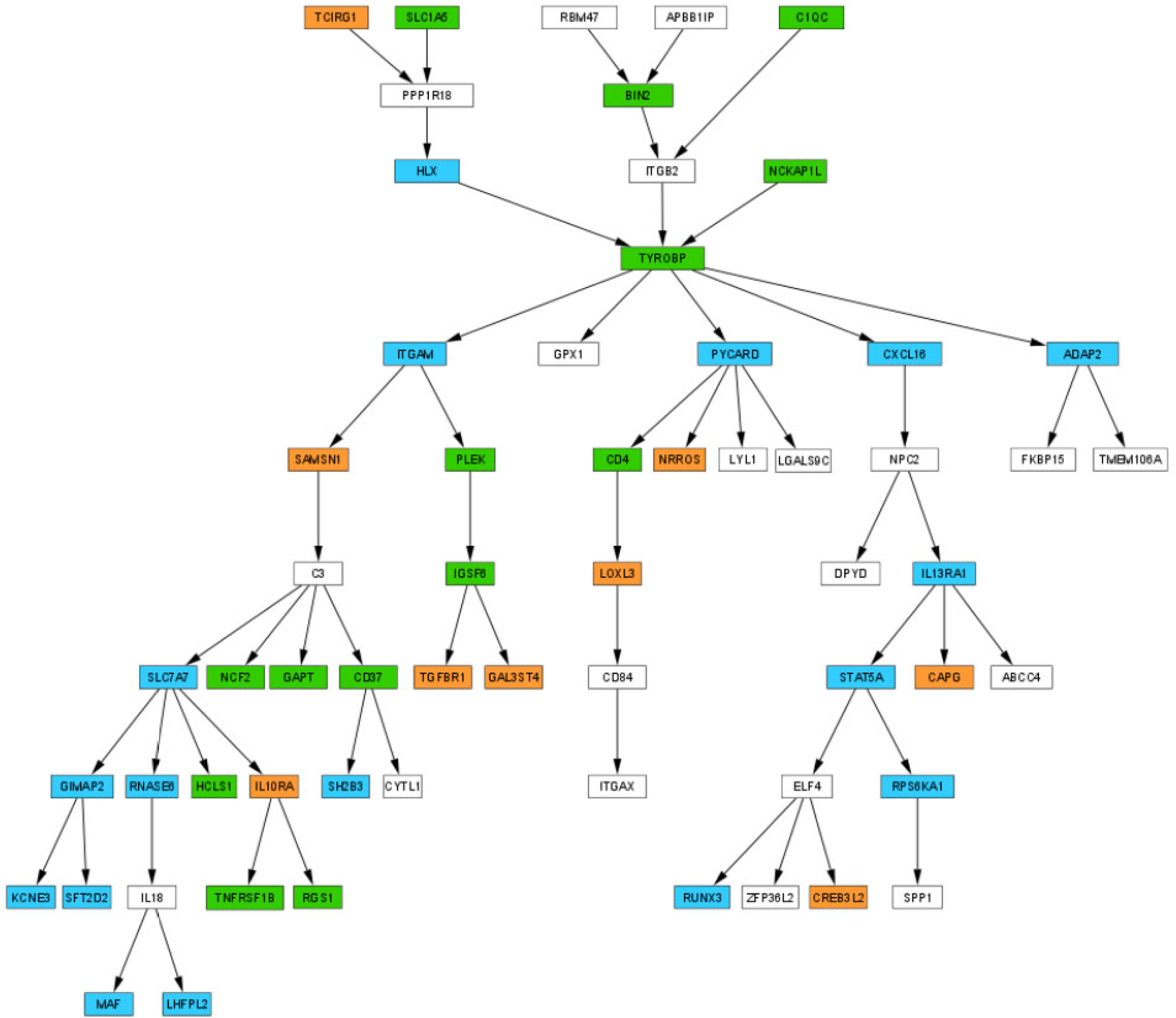
The TYROBP causal network. **(A)** Genes with significant DNA methylation changes in female samples are enriched in the TYROBP causal network (FDR = 0.014). **(B) Core enrichment genes identified by GSEA.** Orange = core enrichment genes identified by GSEA in the analysis of female samples, blue = core enrichment genes identified by GSEA in the analysis of male samples, and green = core enrichment genes identified by GSEA in both male and female samples analysis.

**Supplementary Figure 4.**
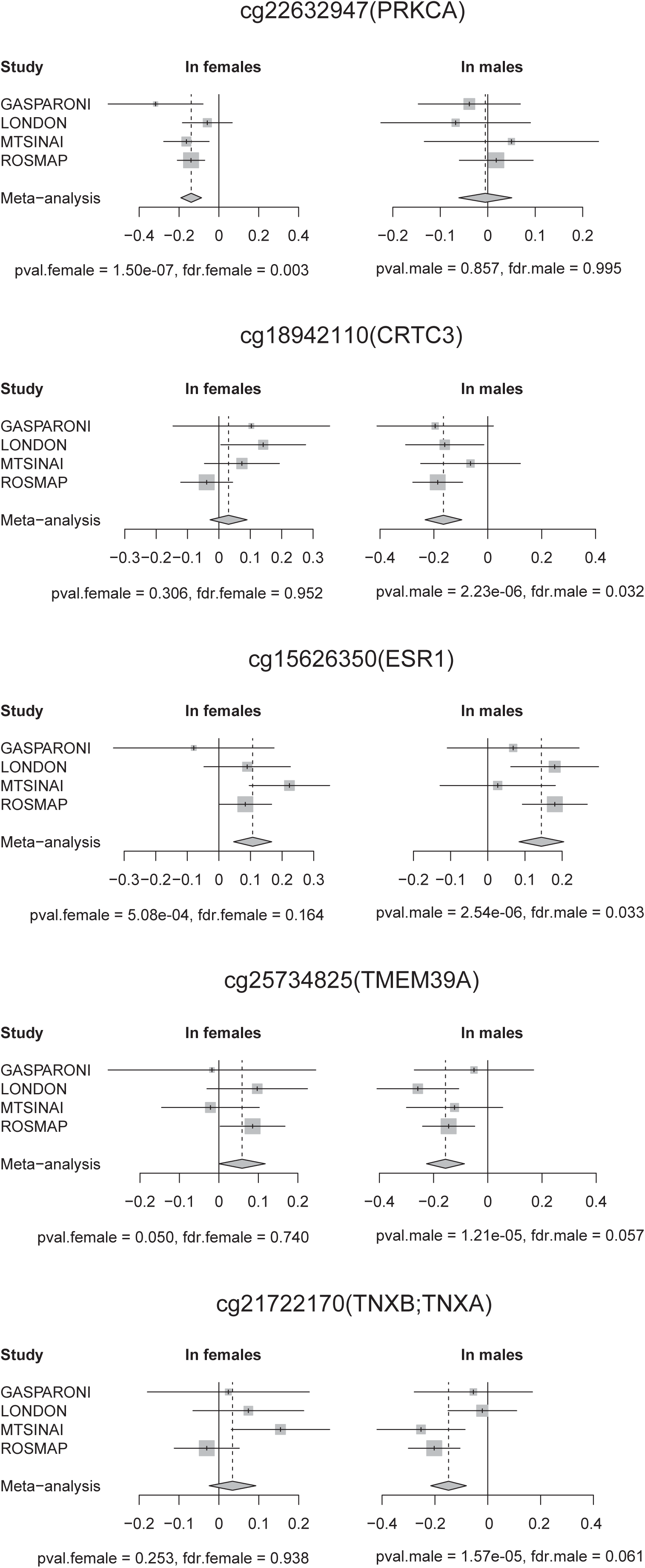
Forest plots for example top CpGs.

